# Treatment advantage in HBV/HIV coinfection compared to HBV monoinfection in a South African cohort

**DOI:** 10.1101/19007963

**Authors:** Tongai G Maponga, Anna L McNaughton, Marije Van Schalkwyk, Susan Hugo, Chikezie Nwankwo, Jantjie Taljaard, Jolynne Mokaya, David A Smith, Cloete van Vuuren, Dominique Goedhals, Shiraaz Gabriel, Monique I Andersson, Wolfgang Preiser, Christo van Rensburg, Philippa C Matthews

## Abstract

**Objective:** Prompted by international targets for elimination of hepatitis B virus (HBV) infection, we performed a cross-sectional observational study of adults with chronic HBV (CHB) infection in South Africa, characterising individuals with HBV monoinfection vs. those coinfected with HBV/HIV, to evaluate the impact of therapy and to guide improvements in clinical care as guidelines for antiviral therapy change over time.

**Design:** We prospectively recruited 115 adults with CHB, over a period of one year at a university hospital in Cape Town, South Africa. HIV coinfection was present in 39 (34%) subjects. We recorded cross-sectional demographic, clinical and laboratory data.

**Results:** Adults with HBV monoinfection were comparable to those with HBV/HIV coinfection in terms of age, sex and body mass. HBeAg-positive status was more common among those with HIV coinfection (p=0.01). However, compared to HBV/HIV coinfection, HBV monoinfected patients were less likely to have had assessment with elastography (p<0.0001) and less likely to be on antiviral treatment (p<0.0001). The HBV monoinfected group was more likely to have detectable HBV viraemia (p=0.04), and features suggesting underlying liver disease including moderate/severe thrombocytopaenia (p=0.007), elevated bilirubin (p=0.004), and APRI score >2 (p=0.02). Three cases of hepatocellular carcinoma were documented, all in patients with HBV monoinfection.

**Conclusion:** In this setting, individuals with HBV monoinfection are disadvantaged in terms of clinical assessment and appropriate antiviral therapy compared to those with HIV coinfection, associated with relatively worse liver health. Enhanced advocacy, education, resources and infrastructure are required to optimise interventions for CHB.

## INTRODUCTION

The burden of chronic infection with hepatitis B virus (HBV) in southern Africa is high, and in this setting the distribution overlaps with populations in which HIV infection is endemic [1]. Due to high rates of coinfection, and the overlap in antiviral regimens, there is a pressing need to develop an enhanced understanding of the interplay between these infections, and to capitalise on opportunities for deploying existing HIV infrastructure to improve HBV diagnosis, clinical care and prevention. International Sustainable Development Goals set a target for elimination of HBV as a public health problem by the year 2030 [2]. Developing insights into the characteristics and outcomes of chronic HBV (CHB) monoinfection and HBV/HIV coinfection will underpin improved strategies for monitoring, prognostication, patient stratification and therapy. Changes in recommendations for combination antiretroviral therapy (cART) regimens for HIV infection should consider the individual and public health impact of therapy on individuals who are co-infected with HBV, particularly in the co-endemic populations of sub-Saharan Africa.

A traditional paradigm suggests that individuals with HBV/HIV coinfection might have a worse prognosis from chronic liver disease than those with HBV monoinfection, typically linked with higher rates of HBeAg-positivity and higher viral loads [1,3], lower CD4+ T cell counts, increased incidence of liver fibrosis [4] and hepatocellular carcinoma (HCC), higher rates of vertical HBV transmission, and higher overall mortality [5]. However, this picture is not consistent between populations (even within individual studies [6]) and the interplay between viruses may be affected by specific characteristics of the host or viral population [7], and by changes in treatment guidelines over time.

Lamivudine (3TC) and tenofovir (most commonly in the form of tenofovir disoproxil fumarate, TDF) are nucleos(t)ide reverse transcriptase inhibitor (NRTI) agents with established track records of safety and tolerability in both HIV and HBV infection. Long-term 3TC use is hampered by a high rate of selection of resistance [8], while TDF is favoured as the genetic barrier to resistance is high [9]. Following publication of the START trial in 2015, HIV treatment is now initiated irrespective of clinical or immunological status [10]. This practice is now enshrined in ‘90-90-90’ targets, seeking to have 90% of cases diagnosed, 90% of these on treatment, and 90% of these virologically suppressed [11].

TDF is one of the recommended choices for the NRTI backbone of first line cART for HIV [12]. In South Africa, second line NRTI combinations include TDF + 3TC, or TDF + FTC, or AZT + 3TC, depending on the patient’s first line regimen [13]. South African ART guidelines recommend HBsAg testing for patients switching from a TDF containing regimen to avoid acute hepatitis flares after TDF withdrawal; in those testing HBsAg positive, TDF is continued as a component of the second line cART regimen. Thus the majority of HIV-positive individuals with HBV coinfection receive treatment with TDF, whether as a component of routine first-line therapy, or incorporated into second-line regimens.

In contrast, for HBV monoinfection, TDF is prescribed only for a subset of patients, based on algorithms that incorporate assessment of the patient (age and sex), virologic status (HBV DNA viral load) and the presence of underlying fibrotic or inflammatory liver disease (ALT, elastography score, ultrasound appearance, biopsy results) [14–16]. WHO targets for 2030 aim for 90% of HBV cases to be diagnosed, and for 80% of those eligible for treatment to be receiving it [17]. However, only a small minority of those living with CHB are currently aware of their status [18], patient stratification and monitoring using laboratory and radiological approaches is not accessible in many settings [18]. Current assessment therefore misses a high proportion of patients who should be treated [19].

Recognising the need for improved characterisation of CHB infection, both alone and in combination with HIV coinfection, we have undertaken a cross-sectional analysis of adults with CHB in an urban cohort in South Africa. We set out to explore the differences between individuals with monoinfection and coinfection, and to compare rates and outcomes of antiviral therapy, with the aim of improving local practice, and with the wider potential to inform future therapeutic guidelines.

## METHODS

### Clinical cohort

We recruited adults with CHB infection, with or without HIV coinfection, attending routine clinical follow-up in hepatology and infectious diseases outpatient clinics at Tygerberg Hospital, a tertiary referral hospital in Cape Town. Healthcare workers at local HIV and primary care clinics within the referral area are encouraged to send all patients diagnosed with HIV-HBV coinfection to the Tygerberg clinic for counselling and baseline investigations including clinical assessment, laboratory tests and elastography. Surrounding clinical centres also refer adults testing HBsAg-positive for assessment and follow-up in the Division of Gastroenterology. Patients are followed at the clinic at intervals according to the baseline findings, typically attending at intervals of six months. Thus, our cohort represents prevalent chronic disease in this community, rather than new incident infections. Cases were defined as being HBsAg-positive (having been under follow-up for a period of >6 months) and were recruited into a cross-sectional cohort (Oxford-South Africa Hepatitis Cohort, ‘OxSA-Hep’), commencing July 2018. We here present the results of a planned interim analysis of data after 12 months of recruitment.

We recorded treatment with antiviral therapy at the time of recruitment to the study, routine clinical laboratory data (including serological markers of HBV infection, creatinine, liver function tests and platelet count), and documented elastography scores when the patient had been assessed by fibroscan. We recorded data in a LabKey database [20] using a unique pseudonymised patient ID number.

### Generation and analysis of laboratory data

Biochemical and serological data were generated on the Cobas 6000 series e601 module analyzer (Roche Diagnostics GmbH, Germany), including alanine aminotransferase (ALT) and aspartate aminotransferase (AST) and serum bilirubin (BR). Platelet counts were obtained using the Advia 2120i analyzer (Siemens Healthcare Diagnostics Inc, USA). HIV and HBV viral loads were measured using Cobas Ampliprep/Taqman tests (Roche Molecular Diagnostics, the Netherlands).

We defined the upper limit of normal (ULN) for ALT as 19 U/L for females and 30 U/L for males [16,21], ULN for AST as 40 U/L, ULN for BR as 17 mmol/L. We calculated liver fibrosis scores, AST to Platelet Ratio Index (APRI) and Fibrosis-4 (FIB-4) as follows:

- APRI = (AST / ULN AST) / (Platelet count × 100)
- FIB-4 = (Age in years × AST) / (Platelet count * √ALT)

We used thresholds of FIB-4 >3.25 (97% specificity, and positive predictive value of 65% for advanced liver fibrosis [22]), and APRI >2 (91% specific for cirrhosis [23,24]).

We measured renal function based on estimated glomerular filtration rate (eGFR) as follows:

- 186 × (creatinine (µmol/L)/88.4) ^-1.154^ × (age in years) ^-0.203^ × (0.742 if female) × (1.210 if black) [25].

Normal renal function is defined as eGFR ≥ 90 ml/min/1.73m^2^. We grouped together individuals with renal impairment, defined as chronic kidney disease (CKD) Stage II (eGFR 60-89 ml/min) or Stage III (eGFR 30-59 ml/min). We defined moderate/severe thrombocytopaenia as a platelet count <75 × 10^3^/μl [26].

### Elastography

Elastography to quantify liver stiffness (an assessment of inflammation and/or fibrosis) is not undertaken consistently in this setting, but is performed in a subset of patients at the discretion of the clinical team using the Fibroscan 402 (Echosens, Paris, France). Elastography scores are not reliable in individuals with a high body mass index or during pregnancy, and would not be undertaken in these circumstances. Due to the cross-sectional nature of the cohort, we recorded elastography scores at a single time point only.

### Treatment indications and outcomes

We have referred to HBV treatment recommendations published by the UK National Institute for Clinical Excellence (NICE) [21], the European Association for the Study of the Liver (EASL) [15], and South African national guidelines [16]. All concur in advising tenofovir or entecavir as first-line options, and recommend that stratification for therapy should be based on laboratory parameters, on the presence and extent of existing liver disease assessed by imaging and/or biopsy, and on consideration of other individual clinical and demographic factors.

Due to resource constraints, liver biopsy is rarely undertaken in this setting and we have primarily used laboratory parameters to assess treatment eligibility. South African guidelines suggest consideration of therapy for individuals who are HBeAg-positive with HBV DNA >20,000 IU/ml and ALT above ULN, or who are HBeAg-negative with HBV DNA >2,000 IU/ml and ALT above ULN, with the aim of achieving durable suppression of HBV DNA to low or undetectable levels and normalisation of ALT [16]. Individuals with HIV coinfection are routinely treated with first line cART that includes HBV-active agents [13]. In practice in this setting, TDF is therefore first line therapy for CHB, both in the presence and absence of HIV infection.

### Statistical analysis

We analysed data using GraphPad prism v 8.0.1. For continuous variables, we used Mann Whitney Test; for categorical variables we used Fisher’s Exact Test. Linear regression was used to assess associations between two continuous variables. For multivariate analysis, we performed multivariate logistic regression using bayesglm function of the R package.

### Ethics

Ethics approval was provided by University of Oxford Tropical Research Ethics Committee (ref. OXTREC 01-18) and Stellenbosch University Human Research Ethics Committee (HREC ref. N17/01/013). All participants provided written valid informed consent. A STROBE statement to support the quality of this observational study is available (Suppl. Table 1).

**Table 1:** Summary of clinical and laboratory parameters recorded from a cohort of 115 adults with HBV infection in Cape Town, South Africa, comparing groups with HBV monoinfection (n=76) versus HIV/HBV coinfection (n=39). For extended version of table, see Suppl data table 3.

|  | Whole Cohort | HBV monoinfection | HIV/HBV coinfection | p-value<sup>a</sup> |
| --- | --- | --- | --- | --- |
| Sex M:F<br>(% male) | 65:50<br>(57%) | 46:30<br>(61%) | 19:20<br>(49%) | 0.24 |
| Median age in years at enrollment<sup>b</sup> (IQR) | 44<br>(36-53) | 45<br>(35-54) | 41<br>(37-50) | 0.51 |
| Median body weight in kg<sup>c</sup> (IQR) | 73<br>(60-84) | 75<br>(60-86) | 71<br>(60-80) | 0.23 |
| Proportion HBeAg positive<sup>d</sup> (%) | 22/107<br>(21%) | 9/70<br>(13%) | 13/37<br>(35%) | **0.011** |
| Proportion with elevated ALT above ULN<sup>e</sup> (%) | 52/113<br>(46%) | 36/74<br>(49%) | 16/39<br>(41%) | 0.55 |
| Proportion with elevated AST above ULN<sup>f</sup> (%) | 31/95<br>(33%) | 20/58<br>(35%) | 11/37<br>(30%) | 0.66 |
| Median BR<sup>g</sup>, mmol/L (IQR) | 7<br>(5-14) | 9<br>(6-17) | 5<br>(4-9) | **0.004** |
| Proportion with elevated BR > ULN<sup>g</sup> (%) | 19/109<br>(17%) | 16/70<br>(23%) | 3/39<br>(8%) | 0.06 |
| Median platelet count<sup>h</sup>, x10<sup>3</sup> / µl (IQR) | 236<br>(163-297) | 227<br>(156-294) | 240<br>(170-307) | 0.41 |
| Proportion with thrombocytopaenia<sup>h</sup> (%) | 11/106<br>(10%) | 11/67<br>(16%) | 0/39<br>(0%) | **0.007** |
| Proportion with APRI score >2 | 8/92<br>(9%) | 8/55<br>(15%) | 0/37<br>(0%) | **0.02** |
| Proportion with FIB-4 score >3.25 | 21/92<br>(23%) | 16/55<br>(29%) | 5/37<br>(14%) | 0.13 |
| Proportion assessed by elastography<sup>i</sup> (%) | 48/115<br>(42%) | 13/76<br>(17%) | 35/39<br>(90%) | **<0.0001\*** |
| Median elastography score, kPa (IQR) | 6.1<br>(4.8-9.0) | 7.8<br>(5.4-10.3) | 5.8<br>(4.5-8.2) | 0.15 |
| Proportion with elastography score >9kPa (%) | 12/48<br>(25%) | 5/13<br>(38%) | 7/35<br>(20%) | 0.26 |
| Proportion with hepatic complications<sup>j</sup> (%) | 20/112<br>(16%) | 15/73<br>(21%) | 5/39<br>(13%) | 0.12 |
| Proportion with CKD stage II or III<sup>k</sup> | 16/111<br>(14%) | 9/72<br>(13%) | 7/39<br>(18%) | 0.79 |

IQR = inter-quartile range; ALT = alanine transferase; ULN = upper limit of normal. <sup>a</sup>p-value for categorical variables by Fisher's Exact Test, and for continuous variables by Mann Whitney test. <sup>b</sup>Age available for 115 individuals. <sup>c</sup>Body weight available for 111/115 individuals. <sup>d</sup>HBeAg status available for 107/115 individuals. <sup>e</sup>ALT available for 113/115 individuals, with ULN defined as 19 iu/ml for females and 30 iu/ml for males. <sup>f</sup>AST available for 95/115 individuals, with ULN defined as 40 iu/ml. <sup>g</sup>BR available for 109/115 individuals; ULN defined as 17 mmol/L. <sup>h</sup>Platelet count available for 106/115 individuals; moderate/severe thrombocytopaenia defined as platelet count <75 x10<sup>3</sup> / ul [26]. <sup>i</sup>Elastography data available for 50/119 individuals. <sup>j</sup>Hepatic complications reviewed for 108/112 individuals, defined as a documented diagnosis of HCC and/or cirrhosis. <sup>k</sup>CKD (chronic kidney disease) stages II and III defined as eGFR <90 ml/min/1.73m<sup>2</sup>. \* also significant on multivariate analysis.

## RESULTS

### Representation of HBV monoinfection and HIV/HBV coinfection in an adult cohort

We recruited 115 adults with HBV infection (57% male), representing 76 adults with HBV monoinfection (66%) and 39 with HBV/HIV coinfection (34%) (Fig 1; Table 1). The full metadata for this cohort are available in Suppl. Table 2. The majority of participants were of South African origin (98/115, 85%; Suppl. Fig 1). There was no significant difference in sex, age or body weight between the monoinfected and coinfected groups (p=0.24, 0.51, and 0.23 respectively; Table 1). HBeAg-positive status was significantly more common among individuals with HIV coinfection (39%) compared to HBV monoinfected individuals (14%); (p=0.01; Table 1; Fig 2A).

**Table 2:** Summary of antiviral therapy treatment and outcomes from a cohort of 115 adults with HBV infection in Cape Town, South Africa, comparing features of those with HBV monoinfection (n=76) versus HIV/HBV coinfection (n=39).

|  | **Whole Cohort** | **HBV monoinfection** | **HIV/HBV coinfection** | **p-value<sup>a</sup>** |
| --- | --- | --- | --- | --- |
| Proportion on antiviral treatment<sup>b</sup> (%) | 72/111 (65%) | 34/72 (47%) | 38/39 (97%) | **<0.0001\*** |
| Median duration of therapy<sup>c</sup>, months (IQR) | 45 (25-77) | 38 (17-69) | 62 (29-81) | 0.11 |
| Median HBV DNA viral load IU/ml<sup>d</sup> (range) | 42 (0-1.7x10<sup>8</sup>) | 86 (0-1.7x10<sup>8</sup>) | 0 (0-1.1x10<sup>8</sup>) | **0.04** |
| Proportion of HBV viraemia undetectable<sup>e</sup> | 41/107 (38%) | 21/69 (30%) | 20/38 (53%) | **0.04** |

<sup>a</sup>p-value for categorical variables by Fisher's Exact Test, and for continuous variables by Mann Whitney test.
<sup>b</sup>Antiviral treatment data available for 111/115 individuals. <sup>c</sup>Duration of therapy defined for 36 HBV monoinfected and 37 HBV/HIV coinfecting individuals. <sup>d</sup>HBV DNA viral load available for 69/76 HBV monoinfected and 38/39 HIV coinfecting individuals. <sup>e</sup>Undetectable HBV DNA defined as viral load in serum below limit of detection of assay (typically HBV DNA <20 IU/ml). \* also significant on multivariate analysis.

**Figure 1:**
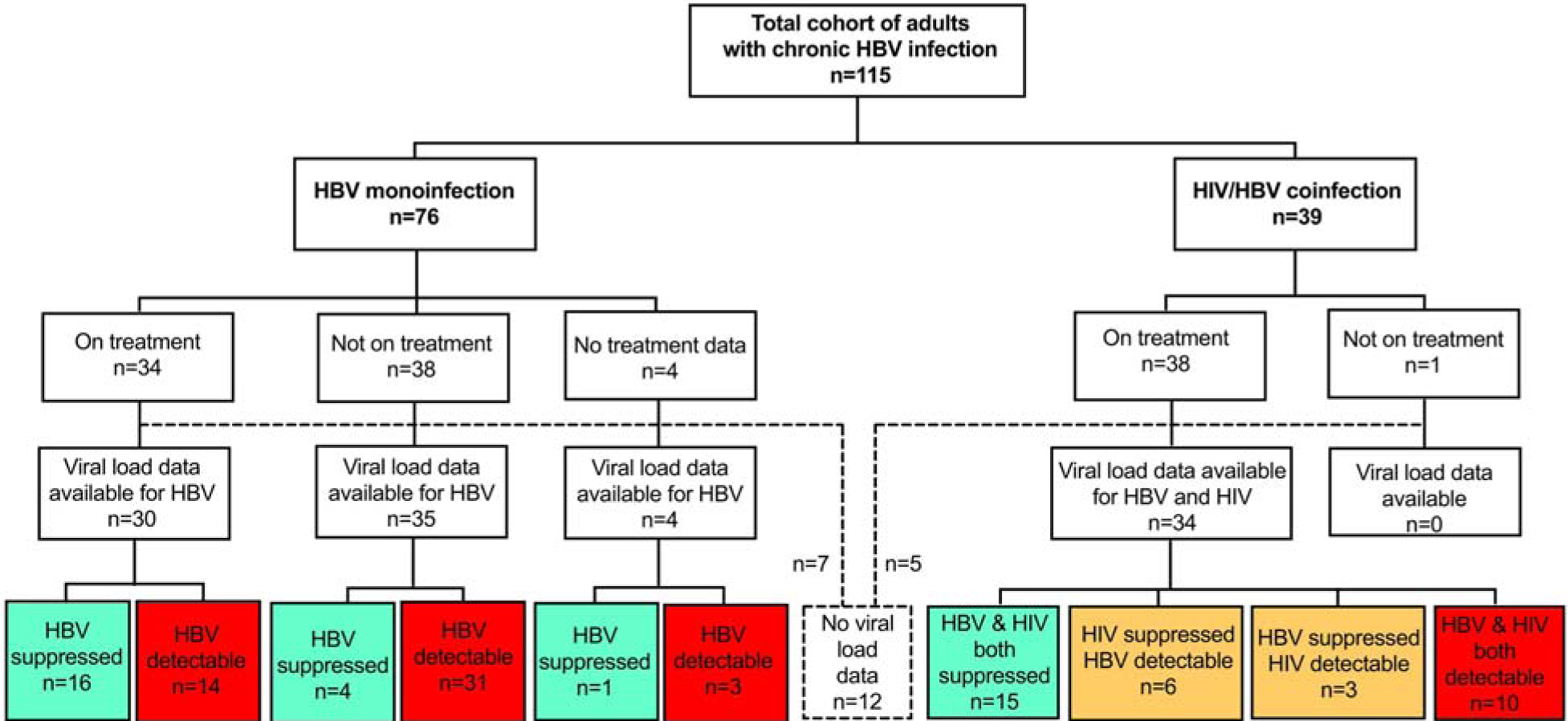
Summary of cohort of 115 adults with HBV infection from Cape Town, South Africa, showing numbers with HBV monoinfection and HIV/HBV coinfection, those receiving therapy and those with suppressed viraemia. Viral load data were not available in 12 cases, of whom 7 were HBV monoinfected and 5 were HBV/HIV coinfected (shown in dashed lines and box). Green boxes indicate number with viraemia suppressed below the limit of detection, yellow indicate number with one virus suppressed and the other detectable, red indicate number with no viraemic suppression.

**Figure 2:**
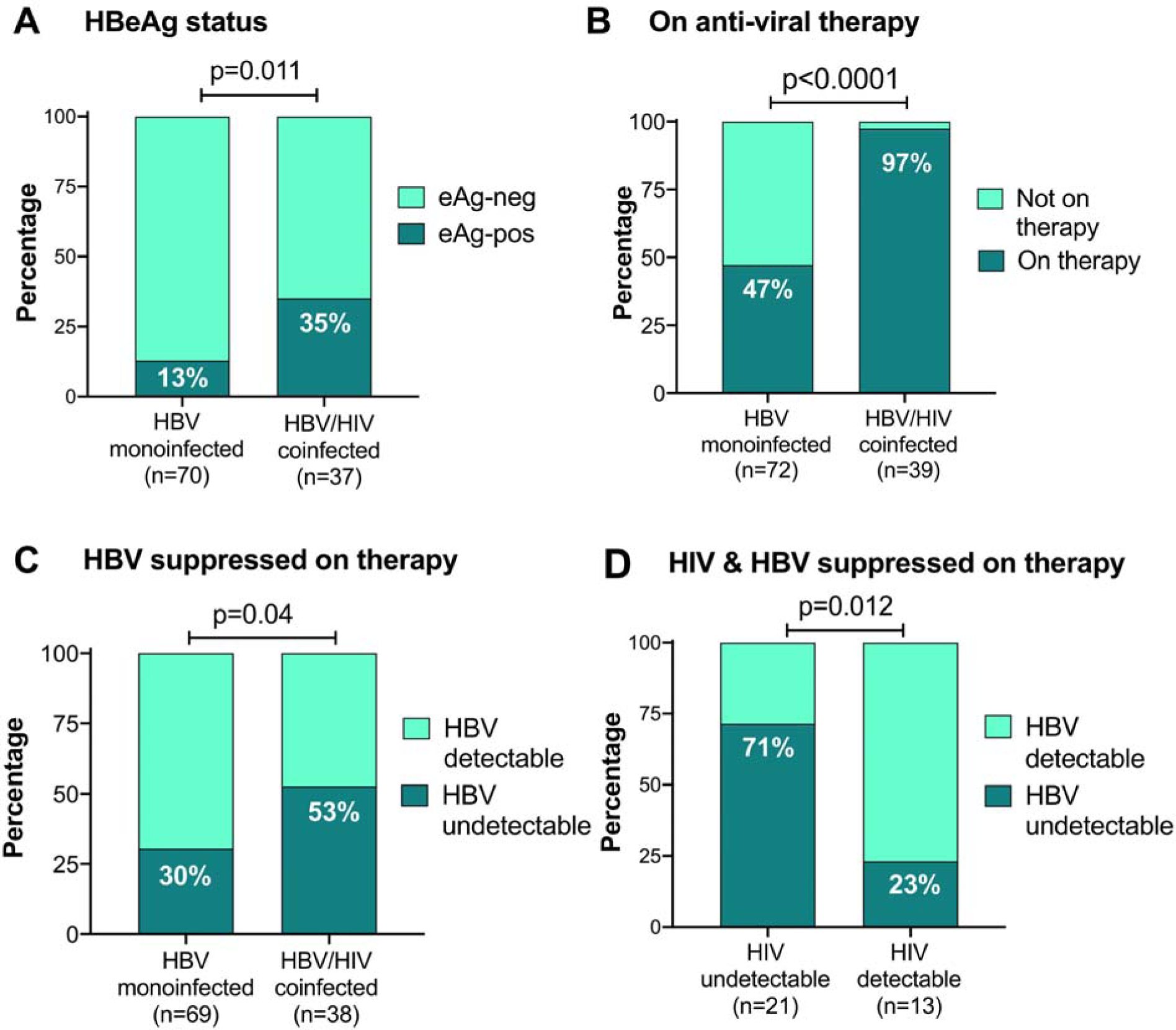
Characteristics of adults with CHB, based on HBeAg status, HBV therapy and virologic response to therapy, comparing those with HBV monoinfection vs HBV/HIV coinfection from a cross-sectional study in Cape Town, South Africa. A: Proportion of each group testing HBeAg-positive. B: Proportion of monoinfected vs coinfected adults receiving antiviral therapy; C: Proportion of monoinfected vs coinfected adults with HBV DNA viral load suppressed below the limits of quantification (<20 IU/ml) on antiviral therapy. D: HBV/HIV co-infected individuals only, showing proportion who have suppressed HIV and/or HBV viral load below the limit of quantification on antiviral therapy. In all cases, p-values calculated by Fisher’s Exact Test, and the number of individuals represented in each group is shown in brackets below the columns.

### Antiviral therapy and virologic outcomes in HBV monoinfection vs HIV/HBV coinfection

Duration of antiviral therapy was recorded for 36 individuals with HBV monoinfection and 37 individuals with HIV coinfection (median 38 vs. 62 months, respectively; p=0.1). As anticipated, based on HIV treatment guidelines [13], a significantly greater proportion of the coinfected group was on antiviral therapy compared to the monoinfected group (97% vs. 47% respectively, p<0.0001; Fig 2B; Table 2). Accordingly, HBV DNA was more likely to be suppressed below the limit of detection in coinfected than monoinfected patients (53% vs 30%, respectively; p=0.04; Fig 2C; Table 2), and absolute values for HBV DNA viral load (VL) were also significantly lower in the coinfected group than the monoinfected (p=0.04; Suppl Fig 2A; Table 2).

In the HBV/HIV coinfected group, 38/39 adults were on antiviral therapy that included HBV-active agents (TDF and/or 3TC); one patient was not on therapy. HIV VL data were available for 35/38, being undetectable in 22/35 (63%), and detectable but <1000 RNA copies/ml in a further 5/35 (14%). Overall, suppression of HIV viraemia was associated with suppression of HBV viraemia (p=0.01, Fig 2D; Suppl Fig 2B). Among individuals with undetectable HIV VL, HBV DNA was still detectable in 6/21 (29%). Among these, one was being treated with 3TC as the only HBV-active agent, with HBV DNA VL 1.0 × 10^5^ IU/ml. Suppression of HIV viraemia suggests adherence to cART, and the high HBV viral load may therefore be consistent with 3TC resistance. The other five were receiving TDF-based combination therapy, making drug resistance a less likely explanation for HBV viraemia (HBV VL 20-5444 IU/ml). Treatment start date was ≥15 months prior to the date of recruitment (recorded in all 6 cases), making it unlikely that HBV viraemia was detectable because therapy had only recently been instituted.

### Proportion of untreated HBV monoinfected patients who meet treatment criteria

We reviewed the data for 38 HBV monoinfected individuals off therapy to determine the proportion of these who met treatment criteria. Using South African guidelines [16], 8/38 (21%) were treatment eligible, comprising individuals with ALT >ULN of whom four were HBeAg-positive with HBV DNA >20,000 IU/ml, and four were HBeAg-negative with HBV DNA >2,000 IU/ml.

### Prevalence of liver disease: laboratory data

We assessed laboratory data and elastography scores as markers for underlying liver disease. There was no significant difference in ALT or AST between monoinfected vs. coinfected individuals (p=0.8 and p=0.6, respectively). For BR, the absolute values were significantly higher in those with monoinfection (p=0.004), and a higher proportion of the population had an elevated value in the moninfected group (23%) vs. the coinfected group (8%), although this difference did not reach statistical significance (p=0.06); Fig 3A,C. Thrombocytopaenia can be an indicator of chronic liver disease as a result of a variety of mechanisms, including splenic sequestration and myelosuppression [26]. Strikingly, all the individuals with moderate/severe thrombocytopaenia were in the HBV monoinfected group, representing 16% of this group vs. 0% in those with coinfection (p=0.007); Fig 3B,D, suggesting a subgroup of individuals with established liver disease.

**Figure 3:**
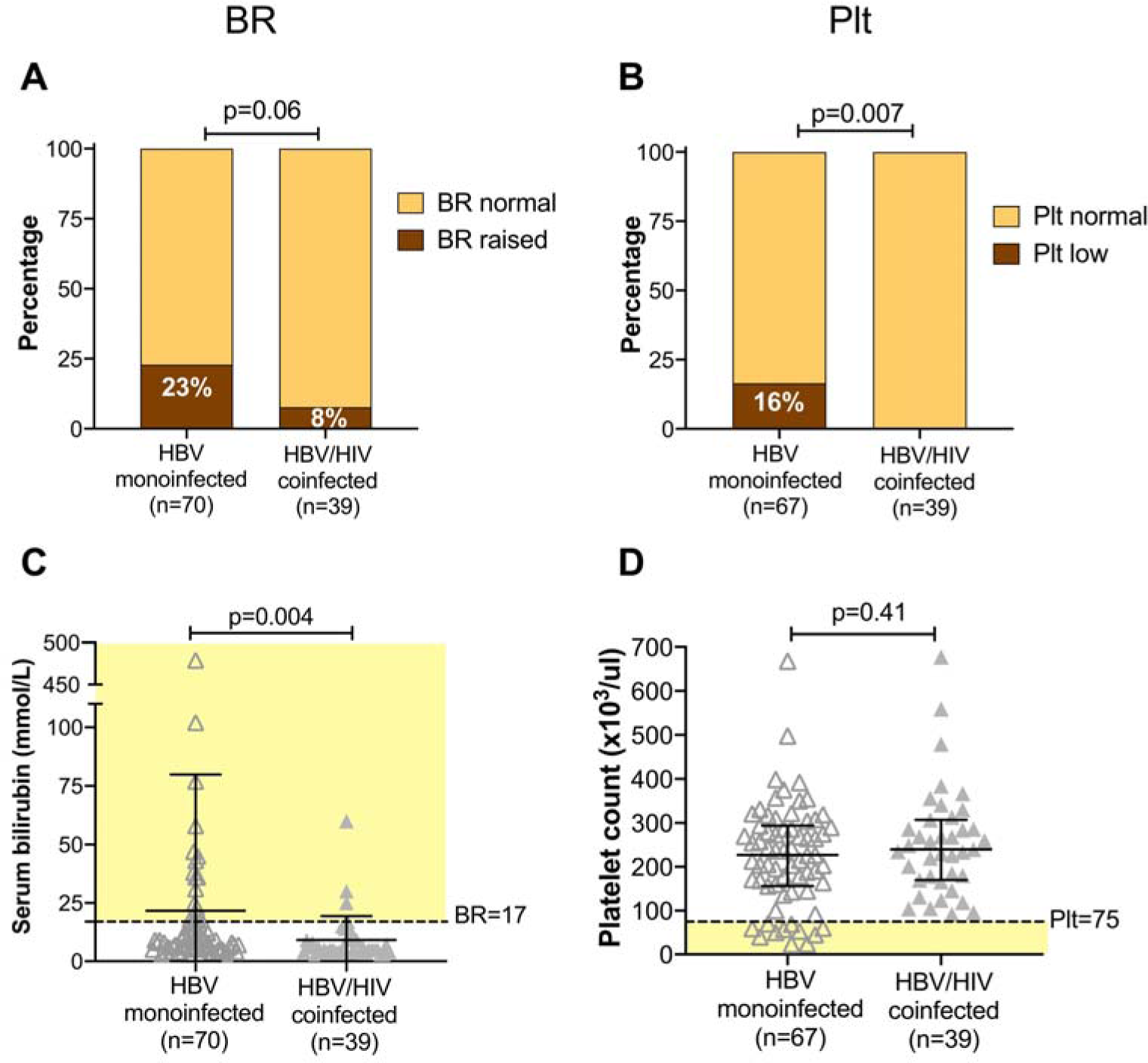
Assessment of liver disease in adults with HBV monoinfection vs HBV/HIV coinfection in a cross-sectional study in Cape Town, South Africa based on serum bilirubin and platelet count. BR - serum bilirubin; Plt - platelet count. Panels A and B: p-values by Fisher’s Exact Test comparing the number in each group falling above and below specified thresholds (BR>17 mmol/L; moderate/severe thrombocytopaenia defined as plt <75 × 10^3^/μl). Number in each group indicated in brackets under the columns; this varies according to data availability. Panels C and D: p-values by Mann Whitney test; bars indicate median and IQR. Pale yellow shading shows population defined as following outside reference range. C: serum bilirubin levels are significantly elevated in the group with HBV monoinfection. D: Distribution of plt highlights all of those with moderate/severe thrombocytopaenia (below dashed line) are in HBV monoinfected group.

We calculated fibrosis scores from laboratory data, finding a significantly higher proportion of the HBV monoinfected group had APRI scores predictive of cirrhosis (p=0.02; Fig 4A). There was a trend in the same direction, although non-significant, for FIB-4 (p=0.1, Fig 4B).

**Figure 4:**
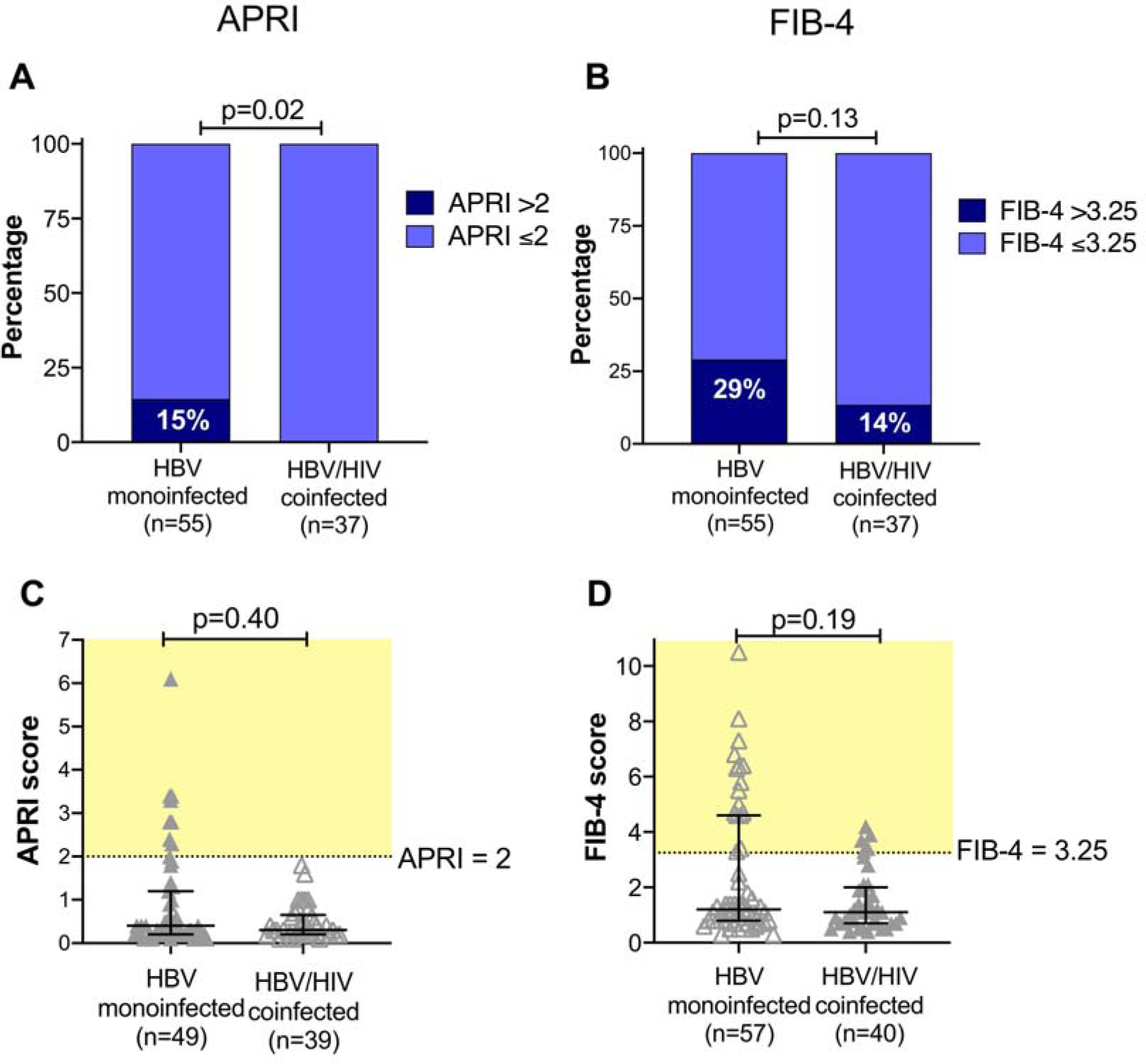
Assessment of liver disease using fibrosis scores derived from laboratory data in adults with HBV monoinfection vs HBV/HIV coinfection in a cross-sectional study of adults in Cape Town, South Africa. APRI - AST to Platelet Ratio Index; FIB-4 - Fibrosis-4 score (for formulae, see methods). Number in each group indicated in brackets under the columns; this varies according to data availability. Panels A and B: p-values by Fisher’s Exact Test comparing the number in each group falling above and below the threshold for cirrhosis/fibrosis (APRI >2; FIB-4 >3.25). Panels C and D: p-values by Mann Whitney test and bars indicate median and IQR. Pale yellow shading shows population defined as falling above the defined threshold for liver disease. No significant difference in median value for either score, but the scatter plots show that individuals with scores above the threshold are predominantly in the mono-infected group.

### Prevalence of liver disease: elastography and clinical data

Due to limited clinical availability, elastography is not routinely undertaken, and data were only available in 48/115 patients (42%). Individuals with HBV/HIV coinfection were more likely to have been assessed by elastography, in whom it was undertaken in 35/39 cases (90%) compared to only 13/76 (17%) of individuals with HBV monoinfection (p<0.0001; Table 1). The median elastography score was higher in the HBV monoinfected group compared to those with coinfection (7.8 kPa vs 5.8 kPa), and a greater proportion of the HBV monoinfected group met the stringent treatment threshold of 6 kPa [21] (9/13 (64%) monoinfected vs. 16/35 (47%) coinfected patients. Neither of these differences reached statistical significance (p=0.2, in both cases), but this comparison is limited by small numbers in the monoinfected group. There was no difference in ALT values between the monoinfected vs coinfected patients with documented fibroscan scores (median ALT 22 vs. 23 U/L, respectively), suggesting that elastography was not systematically undertaken for HBV monoinfected patients with biochemical evidence of inflammatory liver disease.

We reviewed clinical records for complications reported by specialist doctors in 112/115 cases (data not available for three patients). Among these, 20/112 (18%) had hepatic complications of HBV infection, including 18 cases of cirrhosis (16% prevalence) and 3 cases of HCC (3% prevalence); two patients had both cirrhosis and HCC. All HCC cases occurred in the context of HBV monoinfection. The prevalence of hepatic complications was almost twice as high in HBV monoinfection (15/73; 21%) compared to HBV/HIV coinfection (5/39; 13%), although this difference did not reach statistical significance due to small numbers in each group (p=0.1; Table 1).

### Evidence of renal dysfunction

Chronic kidney disease (CKD) can arise in association with HIV infection, and is also a potential side effect of TDF therapy [27]. Therefore, we examined this cohort for the presence of CKD based on calculation of eGFR, available for 111 individuals. Due to small numbers, we grouped together CKD stage II and III, finding a prevalence of 16/111 (14%). There was no difference in the prevalence of CKD between individuals with HBV monoinfection vs. HBV/HIV coinfection (CKD prevalence 9/72 (13%) vs. 7/39 (18%), respectively, p=0.6), and no evidence of nephrotoxicity among those on TDF (CKD present in 9/65 (14%) of treated individuals, vs. 7/43 (16%) of untreated; p=0.8). While we cannot exclude the possibility that selection bias for treatment might mean TDF is more frequently started in individuals with normal renal function, these results are nevertheless reassuring in excluding any significant on-treatment nephrotoxicity.

### Multivariate analysis

Based on small numbers, we did not anticipate multivariate analysis to demonstrate many significant variables, but undergoing assessment by elastography and being prescribed antiviral therapy remained significant on multivariate analysis (Tables 1 and 2, footnotes).

## DISCUSSION

### Context and primary conclusions

Developing a detailed understanding of the characteristics of CHB, and the clinical interplay with co-endemic HIV, is important to drive forward improvements in care provision, with gains to be made both for individuals and populations. Although successful HBV vaccination campaigns are well established, modelling studies predict long timeframes to elimination [28,29], and programmes to enhance diagnosis and treatment will therefore be crucial in the decade ahead.

The traditional paradigm suggests that patients with HBV/HIV coinfection may have worse outcomes than those with HBV monoinfection, and we confirmed a significantly higher prevalence of HBeAg-positive status in those with HIV coinfection, characteristically associated with higher HBV viral loads and acceleration of inflammatory or fibrotic liver disease. However, our data indicate that the monoinfected group is disadvantaged compared to those with coinfection, evidenced by the significant association between monoinfection and lower rates of access to appropriate clinical surveillance, lower rates of antiviral treatment (even among those who meet treatment criteria), and more evidence of liver disease (hyperbilirubinaemia, thrombocytopaenia and elevated APRI). A previous report from the same setting also documents elevated rates of severe fibrosis in HBV monoinfected compared to their HBV/HIV coinfected counterparts [30], although this observation is not consistent as another African study found elevated APRI scores were more likely in patients with coinfection [4].

In our cohort, there was no difference in the prevalence of elevated hepatic transaminases (ALT and AST) between monoinfected and coinfected patient groups. It is important to observe that significant evidence of progressive liver disease can be present even in the absence of abnormalities in liver enzymes. This highlights the lack of sensitivity of liver function tests as biomarkers, and uncertainties around appropriate thresholds for ‘normal’ in different populations highlights the need for clinical surveillance that incorporates other markers.

The worse overall outcomes among the population with HBV monoinfection may stem from the sustained neglect of HBV infection as a clinical and public health problem [18], relatively complex algorithms for HBV treatment stratification, and less consistent and rigorous access to tenofovir monotherapy [31], and may also be influenced by a referral bias. HCC can arise independent of liver fibrosis [16], and therefore even among HBV-infected individuals with no objective evidence of inflammatory or fibrotic liver disease, there is a potential for the emergence of malignancy. Irrespective of the potential bias in the nature of the patients we have been able to assess, our data suggest that adults with HBV monoinfection may be coming to clinical care too late, underlining a pressing need for simplified, scalable approaches to HBV diagnosis as well as treatment [32].

### HBV suppression on therapy

Despite the immunological defect associated with HIV infection and the higher prevalence of HBeAg+ status in this group, HBV viraemia can be successfully suppressed in the majority of coinfected patients using conventional TDF-based cART regimens. However, even on appropriate antiviral therapy, over a quarter of patients still had detectable HBV viraemia. This may be explained by sub-optimal adherence to therapy, pharmacokinetics (drug absorption, levels within liver tissue), or drug resistance, and the long timeframes over which HBV viraemia may decline on therapy [33]. Even on treatment, HBV is more likely to be suppressed in the context of HIV co-infection, perhaps related to better support and education around treatment adherence, or potentially to therapy with more than one HBV active agent (TDF+3TC) in the co-infected group. Careful clinical surveillance is therefore important even in the context of treatment, and there is a need to consider the role of dual HBV therapy and the potential impact of TDF resistance [8,34].

The group of adults with HBV monoinfection whom we assessed according to criteria for therapy points to a burden of infection that is currently untreated, but the patients attending hospital clinics for assessment do not necessarily represent the wider HBV population. Broad community-level surveillance would be required to capture a true picture of the proportion of adults with HBV who meet current treatment criteria. There is also a need for critical appraisal of guidelines, with a view to considering whether thresholds for therapy are appropriate in different settings.

### Changes to cART regimens with influence on HBV therapy

Changes to cART recommendations are developing, based on the success of dual therapy with dolutegravir (DTG) regimens, both in the context of prior viraemic suppression [35] and for salvage after failure of first-line treatment [36,37]. DTG/3TC is a common combination, but other regimens do not contain any HBV active agents, (e.g. DTG combined with rilpivirine (RPV) [38], emtricitabine (FTC) [39], or ritonavir-boosted lopinavir (LPVr) [37]). These regimens have not yet been robustly evaluated in African populations, and are deemed inappropriate in the context of HBV coinfection [40], although there are data demonstrating the efficacy of TDF/FTC in mediating HBV suppression [41]. As dual therapy combinations enter clinical practice in Africa, enhanced awareness and scrutiny will be critical to ensure that HBV-positive individuals are diagnosed and treated with appropriate HBV-active agents, and that there is caution regarding the potential for selection of HBV drug resistance on DTG/3TC treatment.

### Drug resistance

We identified a patient with phenotypic evidence of 3TC-resistant HBV in our cohort, congruent with evidence for the consistent selection of resistance among individuals on 3TC monotherapy [42], and in keeping with another African study that documented a high prevalence of 3TC resistance among coinfected patients [4]. Tenofovir resistance is less well substantiated, and can be difficult to confirm phenotypically due to the slow rate of HBV DNA suppression after institution of therapy [26]; however, there are reports confirming the potential for resistance to this agent (as previously reviewed [8,34]). In the longer term, generation of HBV sequence data will allow us to determine the prevalence of drug resistance mutations, as well as generating insights into the molecular epidemiology and pathogenesis of CHB [7].

### Caveats and limitations

We have studied a small cohort, representing a distinct geographical setting and focusing on clinical care delivery within the specific environment of a tertiary care university hospital, which benefits from enhanced resource and infrastructure compared to many other healthcare settings. Even in this context, missing data are problematic. Assessment with elastography is not consistently deployed, and serological markers are expensive, and not routinely undertaken at each clinic visit. We were unable to determine specific treatment duration for a proportion of the patients studied, adding to uncertainty about exposure to antiviral therapy in HBV and HBV/HIV infected groups. Our data exemplify a ‘real world’ setting and highlight genuine complex day-to-day clinical challenges of managing these patients.

Our study may be influenced by referral bias: patients with more advanced pathology are most likely to be diagnosed and referred for tertiary-level follow-up, potentially leading us to over-estimate liver disease in those with HBV monoinfection. Outside this specific setting, the majority of cases of HBV monoinfection are undiagnosed, and even those with a diagnosis are infrequently under routine surveillance, less likely to be stratified for treatment, and rarely receive a regular supply of appropriate therapy supported by clinical monitoring; on these grounds, our cohort significantly under-represents the true burden of liver disease. However, accepting the referral bias, head-to-head comparison of comparable monoinfected and coinfected groups under clinical surveillance within the same setting consistently suggest a treatment advantage in the coinfected group, and a risk of progression of liver pathology among those with HBV monoinfection.

We have used platelet count as a marker of liver disease, but this is a non-specific approach, as thrombocytopaenia has diverse potential causes. The sensitivity and specificity of APRI and FIB-4 varies between settings, and further evaluation is specifically required in African populations [43]. We were unable to apply other fibrosis scores, such as gamma-glutamyltranspeptidase-to-platelet ratio (GPR) [43,44], as the extended panel of liver function tests required is not routinely collected.

We have taken a cross-sectional view, from which long-term outcomes can only be extrapolated with caution. We have not been able to capture data regarding important influences on HBV and HIV viral loads, including compliance with treatment and the presence of antiviral resistance. HBV genotype data are not available for this cohort, but are known to have a role in disease progression, and may add future insights [7,45].

## Conclusions

High standards of provision of HIV services, with consistent access to diagnosis, surveillance and tenofovir-based treatment, are advantageous in delivering a high standard of clinical care for those with HBV coinfection. The majority of HBV monoinfected patients remain undiagnosed, but - even among those who know their status and are under follow-up - there remains a gap in care provision, rendering them susceptible to long-term liver complications. In order to make progress towards 2030 elimination goals, service providers should capitalise on the existing infrastructure and investment deployed for HIV as both a precedent and a foundation for improved clinical care for HBV infection.

## Data Availability

All data referred to in the manuscript are available as supplementary files (links provided in manuscript).

https://figshare.com/s/756eff7c317ef041cf0e

## ACKNOWLEDGEMENTS

We are grateful to the clinic and laboratory staff in infectious diseases, virology and gastroenterology at Tygerberg Hospital, Cape Town, and to the patients for their participation in this study.

## SUPPLEMENTARY MATERIAL

These data are available on-line: https://figshare.com/s/756eff7c317ef041cf0e. This link will be converted to a permanent open access DOI (10.6084/m9.figshare.9649400) on acceptance of the paper for publication.

**Suppl table 1: STROBE statement** (STrengthening the Reporting of OBservational studies in Epidemiology) to support the quality of this observational cross-sectional study.

**Suppl table 2: Raw data collected for 112 adults with chronic hepatitis B virus infection recruited into a cross-sectional cohort in Cape Town, South Africa**. Information provided in xlsx and csv format.

**Suppl Fig 1: Ethnic origins (birth country) of participants enrolled into a cross-sectional cohort of HBV infection in Cape Town, South Africa**.

**Suppl Fig 2: Plasma viral loads for HBV and HIV in a cross-sectional study of adults in Cape Town, South Africa**. (A) Distribution of HBV DNA viral load in individuals with HBV monoinfection and HBV/HIV coinfection; horizontal lines indicate median with whiskers representing IQR; the numbers shown in brackets under each category report the number of individuals represented; p-values by Mann Whitney U test. (B) Relationship between HIV viral load and HBV viral load in serum, with R^2^ and p values by linear regression based on log_10_ viral load; note the point where HIV and HBV VL=0 represents 18 individuals.

## REFERENCES

1. Matthews PC, Geretti AM, Goulder PJ, Klenerman P. Epidemiology and impact of HIV coinfection with hepatitis B and hepatitis C viruses in Sub-Saharan Africa. J Clin Virol. 2014;61: 20–33.

2. Griggs D, Stafford-Smith M, Gaffney O, Rockstrom J, Ohman MC, Shyamsundar P, et al. Policy: Sustainable development goals for people and planet. Nature. 2013;495: 305–307.

3. van Griensven J, Phirum L, Choun K, Thai S, De Weggheleire A, Lynen L. Hepatitis B and C co-infection among HIV-infected adults while on antiretroviral treatment: long-term survival, CD4 cell count recovery and antiretroviral toxicity in Cambodia. PLoS One. 2014;9: e88552.

4. Ndow G, Gore ML, Shimakawa Y, Suso P, Jatta A, Tamba S, et al. Hepatitis B testing and treatment in HIV patients in The Gambia-Compliance with international guidelines and clinical outcomes. PLoS One. 2017;12: e0179025.

5. Christian B, Fabian E, Macha I, Mpangala S, Thio CL, Ulenga N, et al. Hepatitis B virus coinfection is associated with high early mortality in HIV-infected Tanzanians on antiretroviral therapy. AIDS. 2019;33: 465–473.

6. Matthews PC, Beloukas A, Malik A, Carlson JM, Jooste P, Ogwu A, et al. Prevalence and Characteristics of Hepatitis B Virus (HBV) Coinfection among HIV-Positive Women in South Africa and Botswana. PLoS One. 2015;10: e0134037.

7. McNaughton AL, D’Arienzo V, Ansari MA, Lumley SF, Littlejohn M, Revill P, et al. Insights From Deep Sequencing of the HBV Genome-Unique, Tiny, and Misunderstood. Gastroenterology. 2018. doi:10.1053/j.gastro.2018.07.058

8. Mokaya J, McNaughton AL, Hadley MJ, Beloukas A, Geretti A-M, Goedhals D, et al. A systematic review of hepatitis B virus (HBV) drug and vaccine escape mutations in Africa: A call for urgent action. PLoS Negl Trop Dis. 2018;12: e0006629.

9. Park E-S, Lee AR, Kim DH, Lee J-H, Yoo J-J, Ahn SH, et al. Identification of a quadruple mutation that confers tenofovir resistance in chronic hepatitis B patients. J Hepatol. 2019;70: 1093–1102.

10. INSIGHT START Study Group, Lundgren JD, Babiker AG, Gordin F, Emery S, Grund B, et al. Initiation of Antiretroviral Therapy in Early Asymptomatic HIV Infection. N Engl J Med. 2015;373: 795–807.

11. 90-90-90: treatment for all. [cited 7 Aug 2019]. Available: https://www.unaids.org/en/resources/909090

12. HIV/AIDS Treatment Guidelines. In: AIDSinfo [Internet]. [cited 7 Aug 2019]. Available: https://aidsinfo.nih.gov/guidelines

13. Meintjes G, Moorhouse MA, Carmona S, Davies N, Dlamini S, van Vuuren C, et al. Adult antiretroviral therapy guidelines 2017. South Afr J HIV Med. 2017;18: 776.

14. Overview | Hepatitis B (chronic): diagnosis and management | Guidance | NICE. [cited 7 Aug 2019]. Available: http://nice.org.uk/guidance/cg165

15. European Association for the Study of the Liver. Electronic address, European Association for the Study of the Liver. EASL 2017 Clinical Practice Guidelines on the management of hepatitis B virus infection. J Hepatol. 2017;67: 370–398.

16. Spearman CWN, Sonderup MW, Botha JF, van der Merwe SW, Song E, Kassianides C, et al. South African guideline for the management of chronic hepatitis B: 2013. S Afr Med J. 2013;103: 337–349.

17. Organization WH, Others. Combating hepatitis B and C to reach elimination by 2030: advocacy brief. World Health Organization; 2016. Available: https://apps.who.int/iris/bitstream/handle/10665/206453/WHO_HIV_2016.04_eng.pdf

18. O’Hara GA, McNaughton AL, Maponga T, Jooste P, Ocama P, Chilengi R, et al. Hepatitis B virus infection as a neglected tropical disease. PLoS Negl Trop Dis. 2017;11: e0005842.

19. Aberra H, Desalegn H, Berhe N, Mekasha B, Medhin G, Gundersen SG, et al. The WHO guidelines for chronic hepatitis B fail to detect half of the patients in need of treatment in Ethiopia. J Hepatol. 2019;70: 1065–1071.

20. Data Management and Workflow Software for Research - LabKey. In: LabKey [Internet]. [cited 22 Aug 2019]. Available: https://www.labkey.com/

21. Nice Clinical Guideline, Hepatitis B (chronic): diagnosis and management. NICE Clinical Guideline; 2013.

22. Sterling RK, Lissen E, Clumeck N, Sola R, Correa MC, Montaner J, et al. Development of a simple noninvasive index to predict significant fibrosis in patients with HIV/HCV coinfection. Hepatology. 2006. pp. 1317–1325. doi:10.1002/hep.21178

23. Lin Z-H, Xin Y-N, Dong Q-J, Wang Q, Jiang X-J, Zhan S-H, et al. Performance of the aspartate aminotransferase-to-platelet ratio index for the staging of hepatitis C-related fibrosis: An updated meta-analysis. Hepatology. 2011. pp. 726–736. doi:10.1002/hep.24105

24. Chou R, Wasson N. Blood Tests to Diagnose Fibrosis or Cirrhosis in Patients With Chronic Hepatitis C Virus Infection. Annals of Internal Medicine. 2013. p. 807. doi:10.7326/0003-4819-158-11-201306040-00005

25. Levey AS, Coresh J, Greene T, Stevens LA, Zhang YL, Hendriksen S, et al. Using standardized serum creatinine values in the modification of diet in renal disease study equation for estimating glomerular filtration rate. Ann Intern Med. 2006;145: 247–254.

26. Afdhal N, McHutchison J, Brown R, Jacobson I, Manns M, Poordad F, et al. Thrombocytopenia associated with chronic liver disease. J Hepatol. 2008;48: 1000–1007.

27. Fontana RJ. Side effects of long-term oral antiviral therapy for hepatitis B. Hepatology. 2009;49: S185–95.

28. McNaughton AL, Lourenço J, Hattingh L, Adland E, Daniels S, Van Zyl A, et al. HBV vaccination and PMTCT as elimination tools in the presence of HIV: insights from a clinical cohort and dynamic model. BMC Med. 2019;17: 43.

29. Nayagam S, Thursz M, Sicuri E, Conteh L, Wiktor S, Low-Beer D, et al. Requirements for global elimination of hepatitis B: a modelling study. Lancet Infect Dis. 2016;16: 1399–1408.

30. Maponga TG, Andersson MI, van Rensburg CJ, Arends JE, Taljaard J, Preiser W, et al. HBV and HIV viral load but not microbial translocation or immune activation are associated with liver fibrosis among patients in South Africa. BMC Infect Dis. 2018;18: 214.

31. Maponga TG, Nwankwo C, Matthews PC. Sustainable Development Goals for HBV elimination in South Africa: challenges, progress, and the road ahead. South African Gastroenterology Review. 2019;17: 15–25.

32. Cooke GS, Andrieux-Meyer I, Applegate TL, Atun R, Burry JR, Cheinquer H, et al. Accelerating the elimination of viral hepatitis: a Lancet Gastroenterology & Hepatology Commission. Lancet Gastroenterol Hepatol. 2019;4: 135–184.

33. Price H, Dunn D, Pillay D, Bani-Sadr F, de Vries-Sluijs T, Jain MK, et al. Suppression of HBV by tenofovir in HBV/HIV coinfected patients: a systematic review and meta-analysis. PLoS One. 2013;8: e68152.

34. Mokaya J, McNaughton AL, Bester PA, Goedhals D, Barnes E, Marsden BD, et al. Hepatitis B virus resistance to tenofovir: fact or fiction? A synthesis of the evidence to date. Infectious Diseases (except HIV/AIDS). medRxiv; 2019. doi:10.1101/19009563

35. Baldin G, Ciccullo A, Borghetti A, Di Giambenedetto S. Virological efficacy of dual therapy with lamivudine and dolutegravir in HIV-1-infected virologically suppressed patients: long-term data from clinical practice. J Antimicrob Chemother. 2019. doi:10.1093/jac/dkz009

36. Capetti AF, De Socio GV, Cossu MV, Sterrantino G, Cenderello G, Cattelan A, et al. Durability of dolutegravir plus boosted darunavir as salvage or simplification of salvage regimens in HIV-1 infected, highly treatment-experienced subjects. HIV Clin Trials. 2018;19: 242–248.

37. Aboud M, Kaplan R, Lombaard J, Zhang F, Hidalgo JA, Mamedova E, et al. Dolutegravir versus ritonavir-boosted lopinavir both with dual nucleoside reverse transcriptase inhibitor therapy in adults with HIV-1 infection in whom first-line therapy has failed (DAWNING): an open-label, non-inferiority, phase 3b trial. Lancet Infect Dis. 2019;19: 253–264.

38. Casado JL, Monsalvo M, Fontecha M, Vizcarra P, Rodriguez MA, Vivancos MJ, et al. Dolutegravir plus rilpivirine as dual regimen in virologically suppressed HIV-1 infected patients in a clinical setting. HIV Res Clin Pract. 2019;20: 64–72.

39. Diaco ND, Strickler C, Giezendanner S, Wirz SA, Tarr PE. Systematic De-escalation of Successful Triple Antiretroviral Therapy to Dual Therapy with Dolutegravir plus Emtricitabine or Lamivudine in Swiss HIV-positive Persons. EClinicalMedicine. 2018;6: 21–25.

40. Rossetti B, Montagnani F, De Luca A. Current and emerging two-drug approaches for HIV-1 therapy in ART-naïve and ART-experienced, virologically suppressed patients. Expert Opin Pharmacother. 2018;19: 713–738.

41. Matthews GV, Seaberg EC, Avihingsanon A, Bowden S, Dore GJ, Lewin SR, et al. Patterns and causes of suboptimal response to tenofovir-based therapy in individuals coinfected with HIV and hepatitis B virus. Clin Infect Dis. 2013;56: e87–94.

42. Gu L, Han Y, Li Y, Zhu T, Song X, Huang Y, et al. Emergence of Lamivudine-Resistant HBV during Antiretroviral Therapy Including Lamivudine for Patients Coinfected with HIV and HBV in China. PLoS One. 2015;10: e0134539.

43. O’Hara G, Mokaya J, Hau JP, Downs LO, McNaughton AL, Karabarinde A, et al. Liver function tests and fibrosis scores in a rural population in Africa: estimation of the burden of disease and associated risk factors. Gastroenterology. medRxiv; 2019. doi:10.1101/19000968

44. Lemoine M, Shimakawa Y, Nayagam S, Khalil M, Suso P, Lloyd J, et al. The gamma-glutamyl transpeptidase to platelet ratio (GPR) predicts significant liver fibrosis and cirrhosis in patients with chronic HBV infection in West Africa. Gut. 2016;65: 1369–1376.

45. Kramvis A. Molecular characteristics and clinical relevance of African genotypes and subgenotypes of hepatitis B virus. S Afr Med J. 2018;108: 17–21.

